# Acute myocardial infarction, 90 Cardiovascular proteins and Paroxysmal ventricular tachycardia: a Mendelian randomization study

**DOI:** 10.1101/2024.09.24.24314334

**Authors:** Ji Sun, Yang Wu, Hao-liang Wu, Ying-ying Ji, Xu-Miao Chen, Cheng-Cheng Ji, Su-Hua Wu

**Affiliations:** Department of Cardiology, the First Affiliated Hospital, Sun Yat-Sen University, 58 Zhongshan Road 2, Guangzhou 510080, China; Department of Vascular Surgery, The First Affiliated Hospital of Sun Yat-sen University, 58 Zhongshan Road 2, Guangzhou 510080, China; NHC Key Laboratory of Assisted Circulation (Sun Yat-Sen University), Guangzhou, China

**Keywords:** Acute myocardial infarction, Cardiovascular proteins, Paroxysmal ventricular tachycardia, Leptin, Mendelian randomization

## Abstract

**Background:** Previous studies have shown that acute myocardial infarction (AMI) may be associated with paroxysmal ventricular tachycardia (PVT). However, the causal effect between AMI and PVT, and whether cardiovascular proteins act as a mediator remain unclear.

**Methods:** The genetic data of AMI, 90 Cardiovascular proteins (CVPs) and PVT were obtained from large-scale genome-wide association studies (GWAS). The Mendelian randomization analysis was applied to evaluate the casual association among the AMI, CVPs and PVT. The inverse variance weighting casual effect was regarded as main statistical method. We further investigated whether CVPs take a mediating role in the pathway from AMI to PVT.

**Results:** There were three positive, including ESM-1 (OR 1.139, 95%CI 1.034-1.255, P-FDR=0.014), GAL (OR 1.105,95%CI 1.025-1.191, P-FDR=0.015) and PRL (OR 1.084, 95%CI 1.004-1.170, P-FDR=0.038), and four negative, including FAS (OR 0.928, 95%CI 0.885-0.974, P-FDR=0.009), LEP (OR 0.938, 95%CI 0.891-0.987, P-FDR=0.018), TNF-R1 (OR 0.930, 95%CI 0.883-0.979, P-FDR=0.014) and TNF-R2 (OR 0.928, 95%CI 0.883-0.974, P-FDR=0.009), casual effects between genetic liability of AMI on CVPs. We identified two positive, including IL-27 (OR 1.371, 95%CI 1.010-1.861, P-FDR=0.045) and MMP-12 (OR 1.330, 95%CI 1.035-1.709, P-FDR=0.045), and two negative, including LEP (OR 0.614, 95%CI 0.384-0.981, P-FDR=0.045) and TF (OR 0.714, 95%CI 0.514-0.992, P-FDR=0.045), casual effects between genetic liability of CVPs on PVT. In addition, the AMI played a positive casual effect on PVT (OR 1.736, 95%CI 1.300-2.318, P-FDR<0.001) mediating by LEP with 5.71%.

**Conclusions:** The AMI was casually correlated to PVT, and LEP acted as a mediator in the pathway from AMI to PVT.

**Key Messages:** *What is already known on this topic?:* Previous retrospective and prospective clinical studies have suggested that attention should be paid to the risk of subsequent malignant ventricular arrhythmias, including ventricular tachycardia and ventricular fibrillation, in patients with acute myocardial infarction.

*What this study adds?:* We used Mendelian randomization to demonstrate the genetic relationship between acute myocardial infarction and paroxysmal ventricular tachycardia, and found that leptin mediated the occurrence of paroxysmal ventricular tachycardia in clinical patients with acute myocardial infarction for the first time. In addition, we also found a large number of cardiovascular proteins closely genetically related to acute myocardial infarction or paroxysmal ventricular tachycardia,

*How this study might affect research, practice or policy?:* Our research provides a new insight which means that appropriate serological detection or intervention for patients with acute myocardial infarction can predict or prevent the occurrence of secondary malignant ventricular arrhythmias and reduce the mortality of patients with acute myocardial infarction in the future. Also, the cardiovascular proteins detected by Mendelian randomization analysis are helpful for the diagnosis and prevention of the two.

Graphical Abstract.

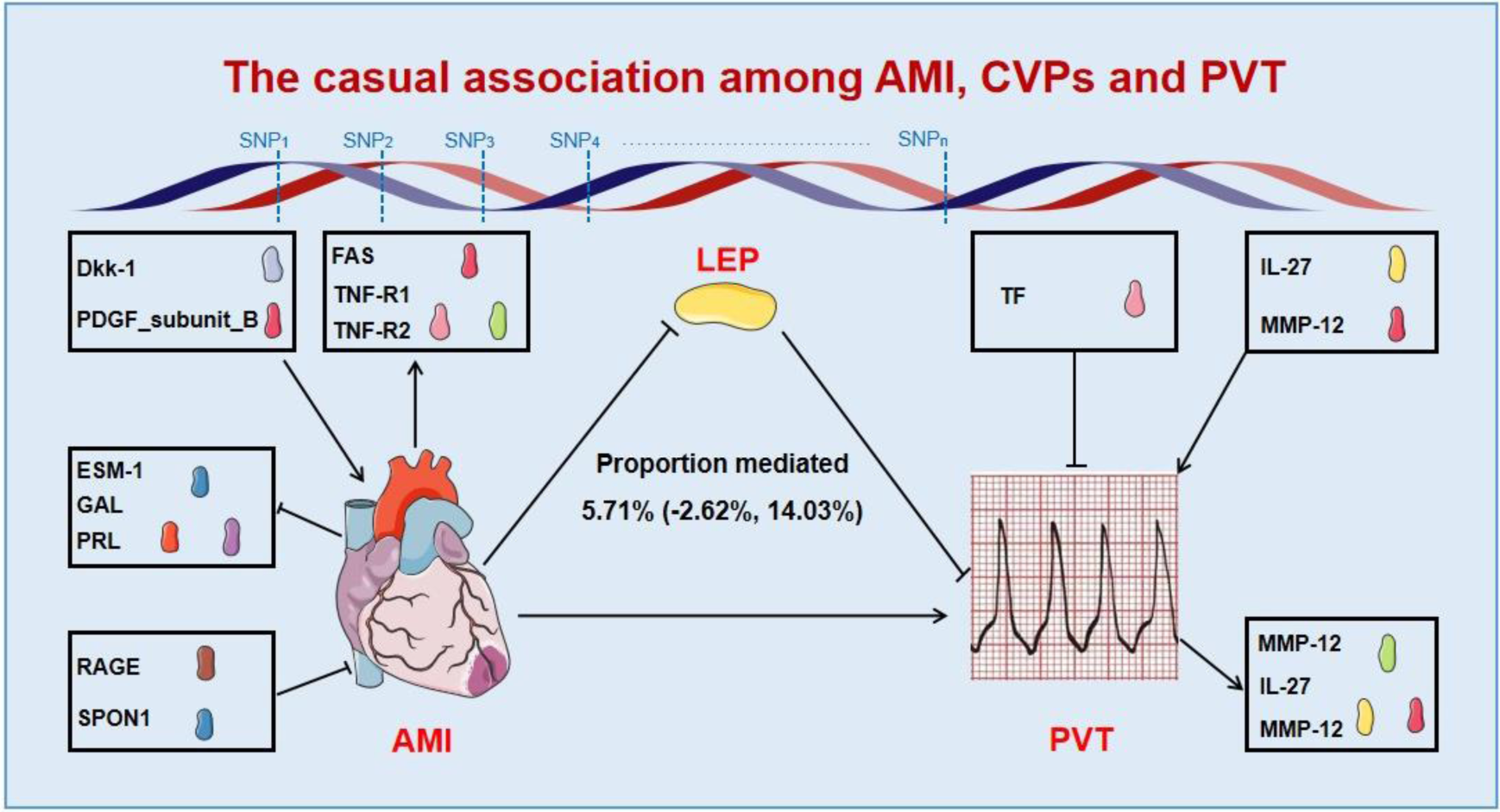

We carefully analyzed the causal relationship among AMI, 90 CVPs and PVT in the complicated MR analysis. We found three positive (ESM-1, GAL and PRL) and four negative casual effects (FAS, LEP, TNF-R1 and TNF-R2) between genetic liability of AMI on CVPs. There were two positive (IL-27 and MMP-12) and two negative (LEP and TF) casual effects between genetic liability of CVPs on PVT. The AMI played a positive casual effect on PVT mediating by LEP with 5.71%. We identified two positive (Dkk-1 and PDGF_subunit_B) and two negative (RAGE and SPON1) casual effects between genetic liability of CVPs on AMI, and thee positive (IL-1ra, MCP-1 and TIE2) casual effects between genetic liability of CVPs on PVT. AMI, acute myocardial infarction; PVT, paroxysmal ventricular tachycardia; CVPs, cardiovascular proteins; AGRP, agouti-related protein; ESM-1, endocan; FAS, tumor necrosis factor receptor superfamily member 6; GAL, galactose-alpha-1,3-galactose; LEP, leptin; PRL, placental growth factor; TNF-R1, tumor necrosis factor receptor 1; TNF-R2, tumor necrosis factor receptor 2; IL-27, interleukin-27; MMP-12, matrix metalloproteinase-12; TF, tissue factor; Dkk-1, dickkopf-related protein 1; PDGF_subunit_B, platelet-derived growth factor subunit B; RAGE, receptor for advanced glycosylation end products; SCF,stem cell factor; SPON1, spondin-1; IL-1ra, interleukin-1 receptor antagonist protein; MCP-1, monocyte chemotactic protein 1; TIE2, angiopoietin-1 receptor.

## Introduction

There is a certain correlation between acute myocardial infarction (AMI) and paroxysmal ventricular tachycardia (PVT). Observational studies showed patients suffered from ventricular arrhythmias, such as premature ventricular beats, ventricular tachycardia and ventricular fibrillation within early-stage after AMI[1]. Moreover, preclinical researches suggested the shortened effective refractory period and delayed conduction of myocardium provide the pathological basis for the occurrence of PVT due to changes in the microenvironment of the damaged myocardium after AMI, like myocardial apoptosis and inflammatory reaction[2, 3]. It is worth noting that PVT appears to act as a mediator for the pathway from AMI to ventricular fibrillation or sudden cardiac death, and that without proper intervention, is often a precursor of ventricular fibrillation and eventually leads to death for patients with AMI. It means that further exploration of the exact mechanism between AMI and PVT could help improve clinical prognosis and life expectancy.

Cardiovascular proteins (CVPs) a group of functional molecules in cardiovascular system play an important role in maintaining the normal structure and function of the heart, regulating the tension and permeability of blood vessels, and affecting the flow and coagulation of blood. Although only one case report suggested that CVPs are closely related to the occurrence of PVT after AMI, a large number of clinical and preclinical studies indicated the significant effect of CVPs on ventricular tachyarrhythmias after AMI[4, 5]. Thus, the function and exact mechanism of CVPs in the pathway from AMI to PVT need to be further explored.

Mendelian randomization (MR), a novel genetic statistical method, is applied to evaluate the causal effect of “exposure factor” on “outcome factor” by extracting genetic variants as instrumental variables (IVs) from Genome-wide association studies (GWAS) which explore the potential associations between different genotypes and phenotypes by testing millions of genetic variants in many individual genomes[6–8]. Moreover, MR analysis possesses a level of evidence comparable randomized controlled trial for randomly assigning genetic variants at the time of conception, so it is affected by confounding or reverse causality hardly[9]. Thus, MR is becoming a popular method in medical research. More critically, no studies have used MR analysis to investigate the association between AMI and PVT, and whether CVPs can act as mediators.

In our study, we launched a comprehensive MR analysis to identify the potential association between AMI and PVT in genetics. Then we dedicated to search possible mediators in the path from a large number of CVPs. At last, we applied reverse causality analysis to explore whether there are other interactions among AMI, CVPs and PVT.

## Materials

### Study design

There are four steps making up our research, as shown in Fig. 1: analysis of causal effects of AMI on PVT (step 1A); analysis of causal effects of AMI on 90 CVPs (step 2A); analysis of causal effects of 90 CVPs on PVT (step 3A) and mediation analysis of CVPs in the pathway from AMI to PVT (step 4). The single-nucleotide polymorphisms (SNPs) was defined as IVs in this study. The MR is based on three core assumptions: (1) the IVs are strongly correlated to the exposure factors; (2) the IVs are not correlated to confounding factors; (3) the IVs do not affect the outcome directly, and it can only affect outcome via the exposure[10].

**Fig. 1.**
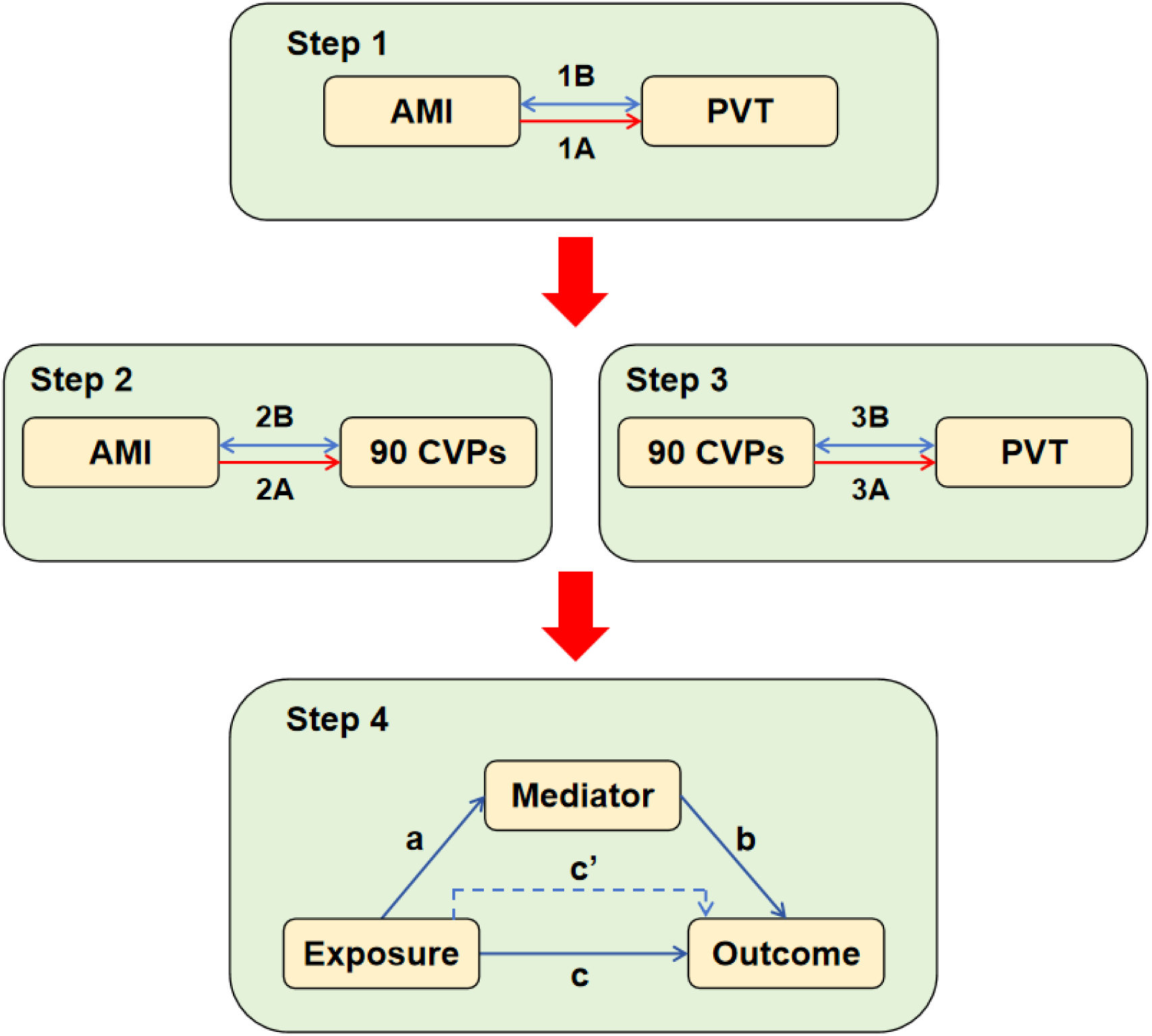
Study overview. Step 1A represents the causal effects of acute myocardial infarction on paroxysmal ventricular tachycardia. Step 1B represents the bi-directional causal effects between acute myocardial infarction and paroxysmal ventricular tachycardia. Step 2A represents the causal effects of acute myocardial infarction on 90 cardiovascular proteins. Step 2B represents the bi-directional causal effects between acute myocardial infarction on 90 cardiovascular proteins. Step 3A represents the causal effects of 90 cardiovascular proteins on paroxysmal ventricular tachycardia. Step 3B represents the bi-directional causal effects between 90 cardiovascular proteins and paroxysmal ventricular tachycardia. Step 4 represents the mediating analysis of cardiovascular proteins in the pathway from the acute myocardial infarction to paroxysmal ventricular tachycardia: path *c* was the total effect of acute myocardial infarction on paroxysmal ventricular tachycardia; path *c’* was the direct effect of acute myocardial infarction on paroxysmal ventricular tachycardia; path *b* was the causal effect of cardiovascular proteins on paroxysmal ventricular tachycardia; path *a* was the causal effect of acute myocardial infarction on cardiovascular proteins. AMI, acute myocardial infarction; PVT, paroxysmal ventricular tachycardia; CVPs, cardiovascular proteins

### Data source

Our genetic data for the AMI was from the latest research, in which 454,787 UK Biobank participants were participating for exome sequencing and analysis, and included in GWAS Catalog database with a study accession GCST90079973 (https://www.ebi.ac.uk/gwas/downloads/summary-statistics)[11]. The genetic data for 90 CVPs came from the previously GWAS analysis included 30,931 individuals[12]. We collected the genetic data for PVT from another GWAS analysis about 456,348 UK Biobank participants, which was also included in GWAS Catalog database with a study accession GCST90043976[13].

There is no new ethical review board approval was required in this study, because of the nature of secondary analysis based on publicly available GWAS summary statistics which was granted with ethical approval and no individual-level data gathering.

### Instrumental variables selection

Firstly, we selected IVs the SNPs significant associating for AMI with a P-value of 5 × 10^-8^. Also, the constant threshold (P ≤ 5 × 10^-8^) was applied to filtrate IVs for 90 CVPs. Afterwards, the SNPs with linkage disequilibrium (LD) meeting the condition r^2^ < 0.001 and distance > 10,000 kb were removed from the IVs[14]. Then, we made a match between exposure and outcome to maintain the consistency of SNP effect on each other. Also, we removed palindromic SNPs, for example an SNP with the A/T or G/C allele, and we got the final IVs. Importantly, there was an appropriate concession (P ≤ 5 × 10^-6^) for part of CVPs to complete subsequent analysis, when few final IVs remain (the sum of final SNPs ≤ 3). In order to minimize the proportion of lost variables, we adjust the P-value reasonably.

The basic information including SNP number, effect allele (EA), other allele (OA), efect allele frequency (Eaf), efect sizes (Beta), standard error (Se), and P-value was extracted successfully. Finally, we further verified the strong correlation between IVs and exposure factors AMI and 90 CVPs by evaluating explained variance (R^2^, R^2^ = [2 × Eaf × (1 - Eaf) × Beta^2^]) and F-statistic parameters (F, F = (Beta/Se)^2^)[15]. The strong association IVs were identified with F-statistic parameters ≥ 10.

### MR analysis

#### Primary analysis

The univariable MR (UVMR) was adopted to assess the causal effects of exposure factor on outcome factor, including AMI on PVT, AMI on 90 CVPs and 90 CVPs on PVT (step 1A, step 2A and step 3A in Fig. 1). A total of five analysis methods were applied in this study containing inverse variance weighted (IVW) random effect, MR egger, simple mode, weighted median and weighted mode, but IVW random effect based on Wald ratios test was the principal method[16]. The odds ratios (ORs) and 95% confidence intervals (CI) were significant presentation pattern of causal effects in our study. When the P-value was no more than 0.05 and the direction was coincided to MR egger according to IVW random effect, the causal effects was considered statistically significant. The false discovery rate (FDR)-corrected P values (P-FDR) calculated with the Benjamini-Hochberg method was regarded as criteria for significance among the significant P-values based on IVW random effect. A P-FDR ≤ 0.05 was considered as significant association; A raw P-value ≤ 0.05 and a P-FDR > 0.05 was considered as suggestive association.

#### Mediation analysis

By the UVMR analysis (step 1A, step 2A and step 3A in Fig. 1), the casual effects of AMI on PVT, AMI on 90 CVPs and 90 CVPs on PVT were verified separately. Then the CVP testified with a casual effect from AMI and a casual effects on PVT was identified as mediator and included in the mediation analysis. Finally, we further evaluated the effect of mediator in the pathway from AMI to PVT (step 4 in Fig. 1).

#### Bi-directional causality analysis

In turn, we regarded PVT as exposure factor and, AMI and 90 CVPs as outcome factor to testify bi-directional causation effect between PVT and PVT, or PVT and 90 CVPs (step 1B and step 3B in Fig. 1). Meanwhile, we regarded 90 CVPs as exposure and 90 CVPs as outcome to evaluate bi-directional causation effect between 90 CVPs and AMI (step 2B in Fig. 1). The method we selected the SNPs significantly associated with PVT was the same as AMI and 90 CVPs.

#### Sensitivity analysis

The heterogeneity of each SNP was assessed by Cochran’s Q test[17]. The potential horizontal pleiotropy effect was evaluated by MR egger regression and MR-PRESSO which was applied further in significant outliers detection and horizontal plural effect revision by excluding outliers[18]. The effect of each SNP on the results was detected Leave-one-out analysis, which operated by removing each SNP in sequence and then performing IVW regression on remaining SNPs to eliminate potential ingredients effect on the estimates[19].

All MR analyses were based on R (v4.3.2) statistical software. The R-based packages including “TwoSampleMR” and “MR_PRESSO”, were used in our MR analysis.

## Results

### Instrumental variable selection

With the threshold of 5 × 10^-8^, we selected 13 SNPs correlated to AMI finally (Additional file 1: Table S2). Meanwhile, we selected 1053 SNPs correlated to 90 CVPs at a level of P < 5 × 10^-8^ or 5 × 10^-6^ totally (Additional file 1: Table S3). A total of 12 SNPs correlated to PVT were identified with a level of P < 5 × 10^-6^ (Additional file 1: Table S4).

### Causal effects among AMI, 90 CVPs and PVT

#### Causal effect of AMI on PVT

We verified that AMI was associated with PVT genetically (Fig. 2A, Additional file 1. Table S5). In Additional file 1. Table S9, there are detail characters of 12 SNPs for PVT. As shown in Fig. 3, The prediction of AMI genetic was correlated with increased the risk of PVT remarkably (OR 1.736, 95%CI 1.300-2.318, P-value < 0.001, P-FDR < 0.001) according to MR analysis.

**Fig. 2.**
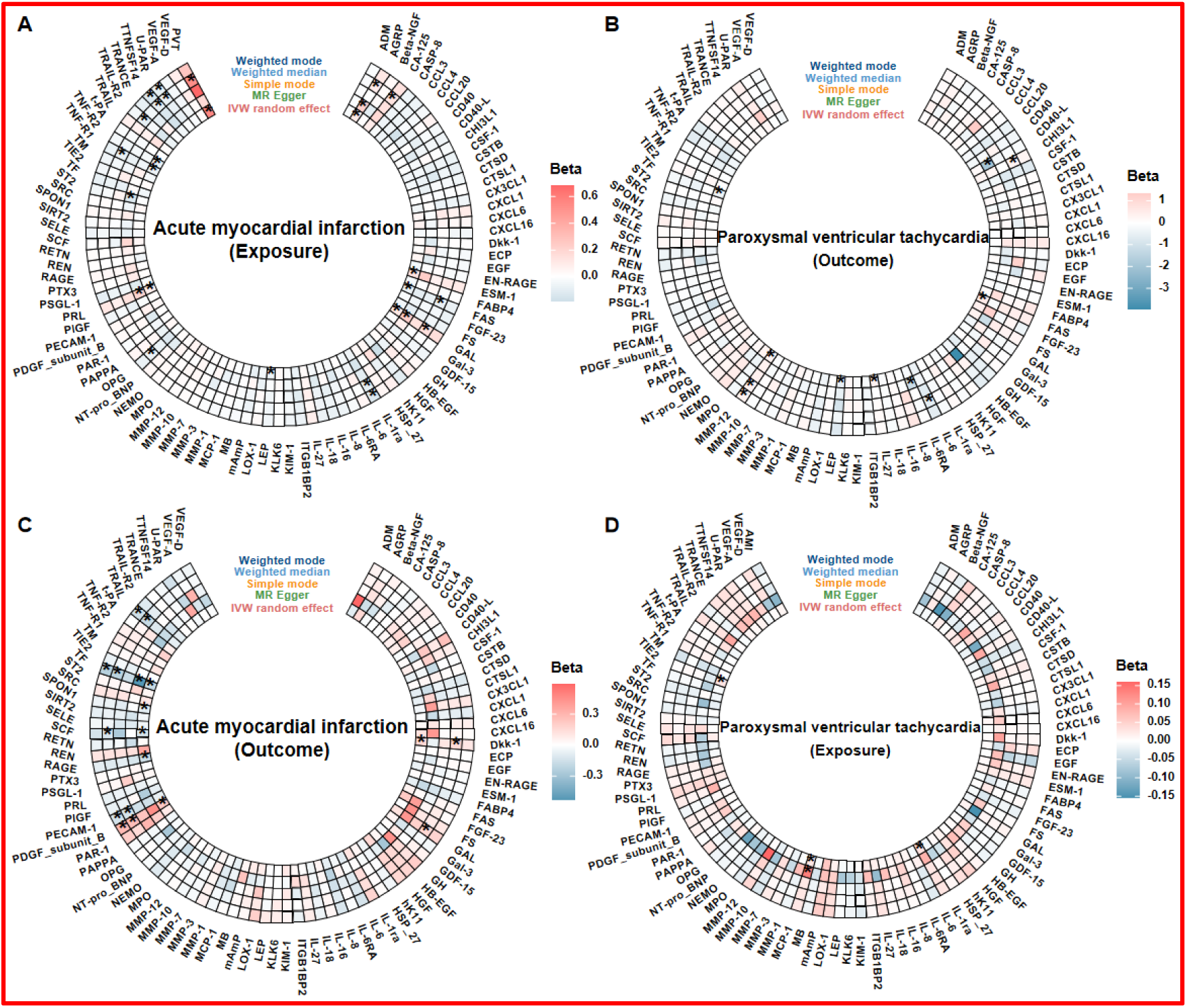
The univariable Mendelian randomization analysis of causal association between acute myocardial infarction, 90 cardiovascular proteins and paroxysmal ventricular tachycardia. A represents the causal effects of acute myocardial infarction on 90 cardiovascular proteins and paroxysmal ventricular tachycardia by five statistical methods. B represents the causal effects of paroxysmal ventricular tachycardia on 90 cardiovascular proteins by five statistical methods. C represents the bi-directional causal effects between paroxysmal ventricular tachycardia and 90 cardiovascular proteins and by five statistical methods. D represents the bi-directional causal effects between paroxysmal ventricular tachycardia and acute myocardial infarction and 90 cardiovascular proteins by five statistical methods. MR, Mendelian randomization; IVW, inverse variance weighted; *, p value ≤ 0.05

**Fig. 3.**
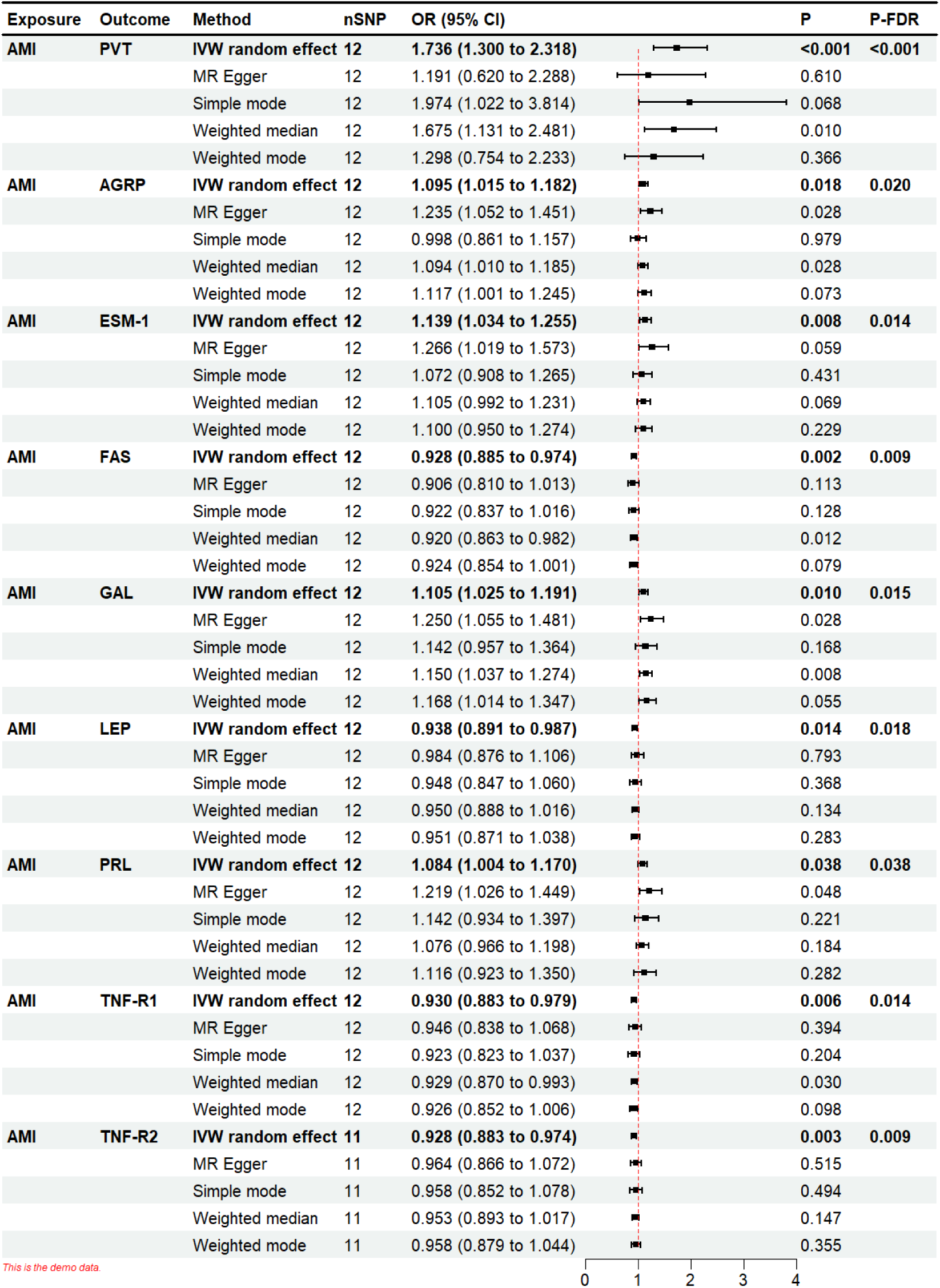
The univariable Mendelian randomization results of causal effects of acute myocardial infarction on paroxysmal ventricular tachycardia and eight cardiovascular proteins. SNP, single nucleotide polymorphism; OR, Odds ratio; CI, confidence interval; FDR, false discovery rate; IVW, inverse variance weighted; AMI, acute myocardial infarction; PVT, paroxysmal ventricular tachycardia; AGRP, agouti-related protein; ESM-1, endocan; FAS, tumor necrosis factor receptor superfamily member 6; GAL, galactose-alpha-1,3-galactose; LEP, leptin; PRL, placental growth factor; TNF-R1, tumor necrosis factor receptor 1; TNF-R2, tumor necrosis factor receptor 2.

**Fig. 4.**
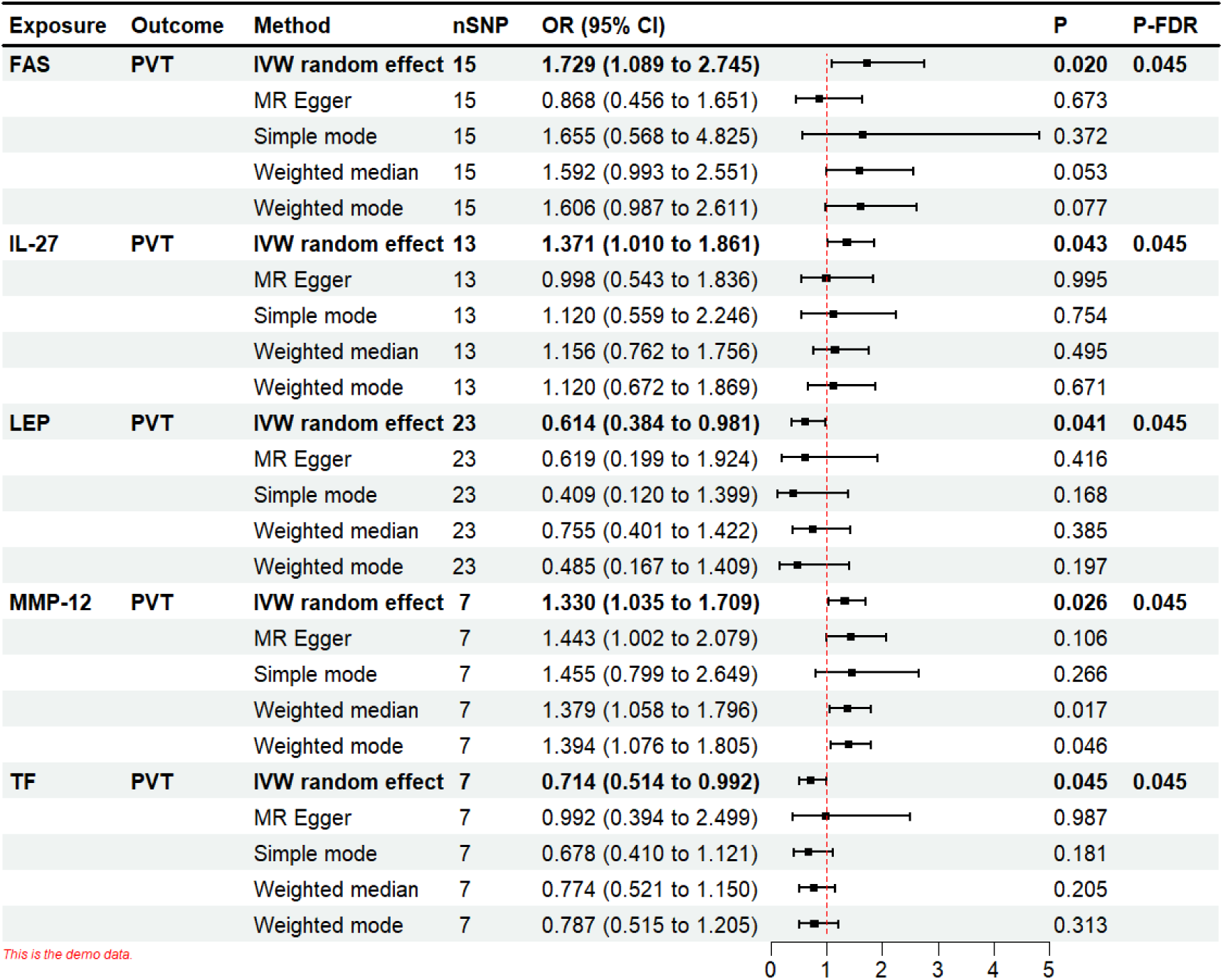
The univariable Mendelian randomization results of causal effects of five cardiovascular proteins on paroxysmal ventricular tachycardia. SNP, single nucleotide polymorphism; OR, Odds ratio; CI, confidence interval; FDR, false discovery rate; IVW, inverse variance weighted; PVT, paroxysmal ventricular tachycardia; FAS, tumor necrosis factor receptor superfamily member 6; IL-27, interleukin-27; LEP, leptin; MMP-12, matrix metalloproteinase-12; TF, tissue factor

#### Causal effect of AMI on 90 CVPs

After multiple UVMR analyses, we verified AMI was associated to 8 CVPs, including AGRP, ESM-1, FAS, GAL, LEP, PRL, TNF-R1 and TNF-R2 (Fig. 2A, Additional file 1. Table S5). The detail characters of SNPs for MR analysis are displayed in Additional file 1. Table S9. The AMI prediction genetic was associated to an increased level of four CVPs AGRP (OR 1.095, 95%CI 1.015-1.182, P-value = 0.018, P-FDR = 0.020), ESM-1 (OR 1.139, 95%CI 1.034 -1.255, P-value = 0.008, P-FDR = 0.014), GAL (OR 1.105, 95%CI 1.025-1.191, P-value = 0.010, P-FDR = 0.015) and PRL (OR 1.084, 95%CI 1.004-1.170, P-value = 0.038, P-FDR = 0.038), which was exhibited in Fig. 3. The occurrence of AMI decreased the level of another four CVPs including FAS (OR 0.928, 95%CI 0.885-0.974, P-value = 0.002, P-FDR = 0.009), LEP (OR 0.938, 95%CI 0.891-0.987, P-value = 0.014, P-FDR = 0.018), TNF-R1 (OR 0.930, 95%CI 0.883-0.979, P-value = 0.006, P-FDR = 0.014) and TNF-R2 (OR 0.928, 95%CI 0.883-0.974, P-value = 0.003, P-FDR = 0.009), shown in Fig. 3.

#### Causal effect of 90 CVPs on PVT

There was a total of five CVPs (FAS, IL-27, LEP, MMP-12 and TF), which were identified associated with PVT, shown in Fig. 2B and Additional file 1. Table S6. The characters of SNPs for 90 CVPs were displayed elaborately (Additional file 1. Table S10). It is shown that FAS (OR 1.729, 95%CI 1.089-2.745, P-value = 0.020, P-FDR = 0.045), IL-27 (OR 1.371, 95%CI 1.010-1.861, P-value = 0.043, P-FDR = 0.045) and MMP-12 (OR 1.330, 95%CI 1.035-1.709, P-value = 0.026, P-FDR = 0.045) are significantly associated with the incidence increased of PVT but LEP (OR 0.614, 95%CI 0.384-0.981, P-value = 0.041, P-FDR = 0.045) and TF (OR 0.714, 95%CI 0.514-0.992, P-value = 0.045, P-FDR = 0.045) are correlated with the decreased risk of PVT in Fig. 3.

### Bi-directional causal effects among AMI, 90 CVPs and PVT

#### Bi-directional causal effect between AMI and PVT

There is no genetic reverse effect between AMI and PVT according to MR analysis with an exchange of exposure and outcome (Fig. 2D, Additional file 1. Table S8). In Additional file 1. Table S12, there are detail characters of 10 SNPs for AMI.

#### Bi-directional causal effect between AMI and 90 CVPs

Although there was no reverse effect between AMI and previous eight CVPs (AGRP, ESM-1, FAS, GAL, LEP, PRL, TNF-R1, TNF-R2), we found another six CVPs were correlated with AMI (Fig. 2D and Additional file 1. Table S7). The detail characters of SNPs for 90 CVPs were displayed in Additional file 1. Table S11. The Dkk-1 (OR 1.167, 95%CI 1.034-1.317, P-value = 0.013, P-FDR = 0.026) and PDGF_subunit_B (OR 1.249, 95%CI 1.095-1.424, P-value = 0.001, P-FDR = 0.006) were significantly correlated with recurrence of AMI, shown in Additional file 2. Figure S1. The others were associated to decrease the risk of AMI extremely including RAGE (OR 0.851, 95%CI 0.730-0.991, P-value = 0.038, P-FDR = 0.040), SCF (OR 0.879, 95%CI 0.778-0.994, P-value = 0.040, P-FDR = 0.040), SPON1 (OR 0.917, 95%CI 0.849-0.990, P-value = 0.026, P-FDR = 0.039) and TF (OR 0.832, 95%CI 0.739-0.936, P-value = 0.002, P-FDR = 0.006), as exhibited in Additional file 2. Figure S1.

#### Bi-directional causal effect between 90 CVPs and PVT

There was no reverse effect between five CVPs (FAS, IL-27, LEP, MMP-12 and TF) and PVT, but we identified the genetic prediction of PVT was correlated with another three CVPs IL-1ra, MCP-1 and TIE2 (Fig. 2C and Additional file 1. Table S8). The characters of SNPs for MR analysis were displayed elaborately in Additional file 1. Table S12. The PVT was associated with increased level of the three CVPs IL-1ra (OR 1.031, 95%CI 1.006-1.057, P-value = 0.017, P-FDR = 0.027), MCP-1 (OR 1.027, 95%CI 1.005-1.049, P-value = 0.018, P-FDR = 0.027) and TIE2 (OR 1.028, 95%CI 1.002-1.054, P-value = 0.032, P-FDR = 0.032), as shown in Additional file 2. Fig S2.

### Sensitivity analyses

There was no genetic pleiotropy biased the results on the basis of MR Egger regression intercept approach with a P-value > 0.05 except FAS on PVT and TF on AMI (Additional file 1: Table S13-16). There was no horizontal pleiotropy according to MRPRESSO analysis and no significant heterogeneity according to Cochran’s Q tests with a P-value > 0.05 except AMI on AGRP and SCF on AMI (Additional file 1: Table S13-16). The UVMR results were verified be credible based on “leave-one-out” analysis, because scale of axis X=0 is not included in the total CI of the SNPs (Additional file 2: Figure S3-6). The scatter plots revealed overall effect of AMI on PVT and 8 CVPs (Additional file 2: Figure S7), 5 CVPs on PVT (Additional file 2: Figure S8), 6 CVPs on AMI (Additional file 2: Figure S8) and PVT on 3 CVPs (Additional file 2: Figure S10). Also, the corresponding forest plots manifesting causal relations were shown in Additional file 2: Figure S11-14.

### Mediation analysis

On the first step, we verified that genetic association of AMI on the recurrence of PVT. Thereafter, we identified eight, AGRP, ESM-1, FAS, GAL, LEP, PRL, TNF-R1 and TNF-R2 from 90 CVPs influenced significantly by AMI. Moreover, from the same 90 proteins, we found five, FAS, IL-27, LEP, MMP-12 and TF which take a casual effect on PVT. Thus, FAS and LEP were taken as potential factors played a mediating effect in the pathway from AMI to PVT. Because of the pleiotropy in the casual effect of FAS on PVT, LEP was identified as unique mediator. The mediation analysis result of LEP in the pathway from AMI to PVT was shown in Fig 5 and Additional file 1: Table S17.

**Fig. 5.**
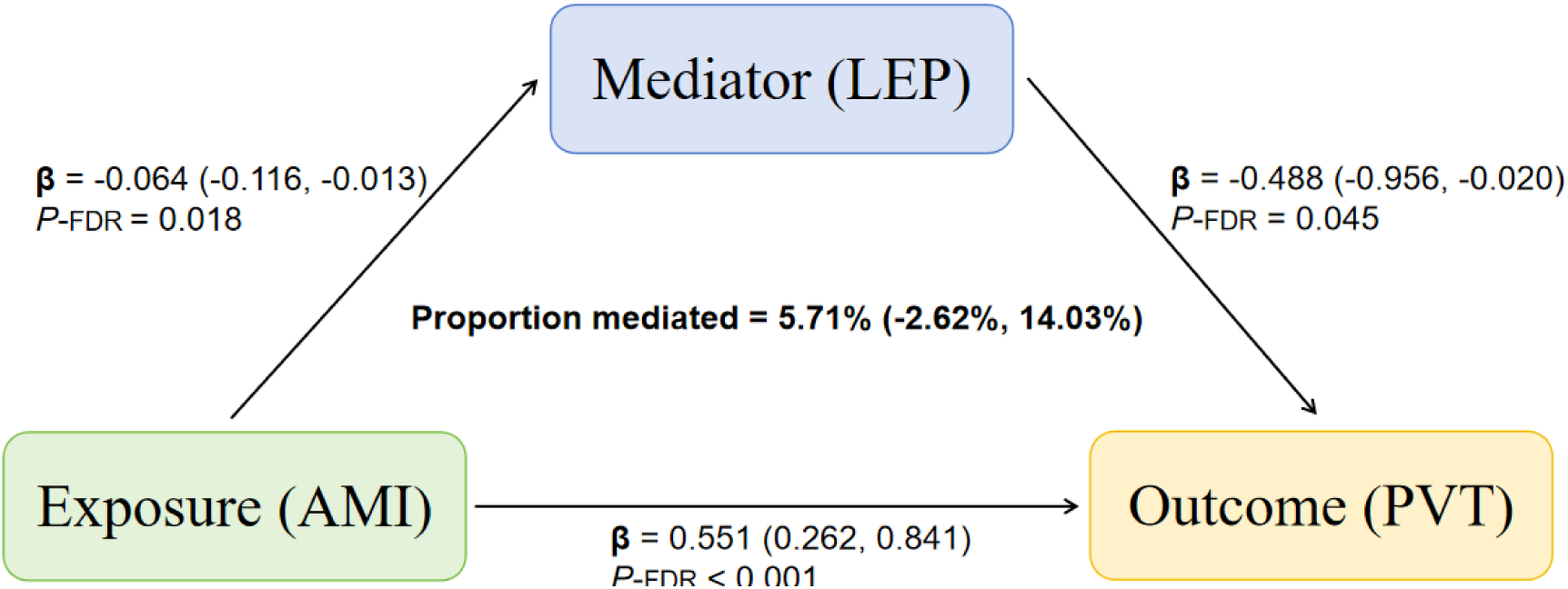
The potential causal evidence summarized from the MR analysis. AMI, acute myocardial infarction; PVT, paroxysmal ventricular tachycardia; LEP, leptin; FDR, false discovery rate.

## Discussion

### Principal findings

In this comprehensive MR analysis, we carefully analyzed the causal relationship among AMI, 90 CVPs and PVT (**Graphical Abstract**). First, we provided the genetic basis for the occurrence of PVT after AMI for the first time. Then, we identified LEP from 90 CVPs as a mediator with 5.71% in the pathway of AMI to PVT by MR mediation analysis.

Moreover, we discovered a large number of proteins from 90 CVPs genetically associated with AMI or PVT, which may contribute to the auxiliary diagnosis or the timely warning before the occurrence of diseases. Specifically, our study indicated seven CVPs (ESM-1, FAS, GAL, LEP, PRL, TNF-R1 and TNF-R2) influenced significantly by AMI, three CVPs (IL-1ra, MCP-1 and TIE2) influenced significantly by PVT, four CVPs (Dkk-1, PDGF_subunit_B, RAGE and SPON1) influencing the onset risk of AMI and four CVPs (IL-27, LEP, MMP-12 and TF) influencing the onset risk of PVT.

### The association between AMI and PVT

The AMI and PVT, two major emergencies of the cardiovascular system, can lead to poor prognosis and even sudden cardiac death, if the treatment measures taken after the occurrence are inappropriate. Previous observational studies and retrospective cohort studies suggested patients with AMI suffered from PVT constantly with poor explanation of causation[1]. There seems to be few evidence proving the potential association between AMI and PVT from randomized controlled study. Some preclinical researches indicated the potential mechanism of PVT caused by AMI, such as shortened effective refractory period and delayed conduction of myocardium attributed to myocardial apoptosis and inflammatory reaction after AMI, in animal models[2, 3]. However, it needs to be explored further for species difference and controllable or uncontrollable factors in animal experiment. The MR analysis is a novel genetic epidemiological statistical method and provides substantial evidence of which the level is comparable to randomized controlled study for genetic variants determined at the time of conception and less susceptible to environmental confounding factors or reverse causality are used for analysis[6–9]. The present MR study indicated AMI took a genetic prediction of the increased risk of PVT but there was no casual effect of PVT on AMI, which was consistent with the conclusion of the current observational research. Thus, our results provided a genetic basis for the potential association between AMI and PVT for the first time. Moreover, our study also cleverly avoids the biases that arise from species differences and controllable or uncontrollable factors in basic experiments by using human genetic data directly, albeit for European populations.

### The mediation effect of CVPs in the association between AMI and PVT

The CVPs generally maintain the normal structure and function of the heart, regulate the tension and permeability of blood vessels, and affect the flow and coagulation of blood in cardiovascular system. Previous case report indicated CVPs were closely related to the occurrence of PVT after AMI without mechanism explanation[4]. Other preclinical studies suggested the significant role of CVPs on ventricular tachyarrhythmias after AMI at the animal model level[5, 20]. In order to identify potential intermediary mediating the occurrence of PVT after AMI, we performed UVMR analysis for three times, between AMI and PVT, AMI and CVPs, and CVPs and PVT. For a result, we testified the casual effect of AMI on PVT, AMI on seven CVPs and four CVPs on PVT after sensitivity analysis. Interestingly, in the later two analyses we discovered that there a common CVP the LEP which can be regarded as the mediator. The mediation analysis shown that genetic prediction of AMI was associated to decreased level of LEP; genetic prediction of LEP was associated with decreased occurrence of PVT and the proportion mediated of LEP was 5.71%. The role of LEP in cardiovascular disease is controversial. Currently, LEP was known as an adipokine that plays an important role in lipid metabolism which was widely researched[21]. High level of LEP was associated with unfavorable outcomes in cardiovascular disorders, but it can elicit cardio-protective effects by reducing cardiomyocyte hypertrophy and apoptosis[22, 23]. Moreover, there are a large number of LEP receptors distributed in the cardiovascular system which provide structural support for LEP regulation of heart rhythm, myocardial electrical homeostasis and vasoconstriction[24]. Our MR result suggested the genetic prediction of AMI was correlated to decreased level of LEP. But part of observational researches indicated the high level of LEP was associated with increased risk of AMI[22]. It did not indicate a causal relationship between the occurrence of AMI and increased LEP level, and the clinically observed increase in LEP level after AMI may be attributed to the treatment or body compensatory regulation. And a prospective cohort study found elevated LEP level after AMI reperfusion therapy[25]. Furthermore, we testified a genetic prediction for lower level of LEP on paroxysm of PVT. A preclinical study in Beagle dogs showed that injection of LEP into the cardiac autonomic nervous system increased the incidence of multiple ventricular arrhythmias after AMI[20]. However, another vitro pre-clinical studies suggested that high-concentration leptin can directly increase QT intervals and trigger ventricular arrhythmias without underlying ionic mechanisms[26]. In brief, these previous studies provide some theoretical basis for our conclusion, and our results also provide new evidence for the role of LEP in occurrence of PVT after AMI. The exact mechanism of LEP mediating PVT after AMI remains to be further explored.

### The association between CVPs and AMI or PVT

For the association between CVPs and AMI, we firstly evaluated the casual effect of AMI on 90 CVPs. Our results suggested AMI prediction genetic was associated to an increased level of ESM-1, GAL and PRL and a decreased level of FAS, LEP, TNF-R1 and TNF-R2. Also, we further assessed the casual effect of 90 CVPs on AMI. we found Dkk-1 and PDGF_subunit_B were significantly correlated with recurrence of AMI, and RAGE and SPON1 were associated to decrease the risk of AMI. Moreover, at the same time we evaluated the casual effect of PVT on 90 CVPs and 90 CVPs on PVT. Our results indicated that IL-27 and MMP-12 took a genetic prediction of increased risk of PVT, and LEP and TF was associated with decreased risk of PVT, and also suggested occurrence of PVT took a casual effect on increased levels of IL-1ra, MCP-1 and TIE2. Interestingly, previous clinical and preclinical study indicated the most of potential associations above[27–30]. But there are some novel CVPs identified correlated with AMI or PVT rarely, like GAL, PRL and TIE2. In conclusion, we provided genetic evidence for casual effect between a great number of CVPs, which can be helpful for auxiliary diagnosis and risk assessment of AMI or PVT. Of course, the underlying mechanism needs to be further explored in subsequent studies.

### Strengths and limitations

To our knowledge, it is the first time to explore the potential association among AMI, a large number of CVPs and PVT with a novel tool MR analysis. Moreover, we provided evidence on the occurrence of PVT after AMI and the potential mechanism through the decreased level of LEP in genetics. Furthermore, we identified seven proteins (ESM-1, GAL, PRL, FAS, LEP, TNF-R1 and TNF-R2) and four (IL-27, MMP-12, LEP and TF) took a casual effect of AMI and PVT respectively which may be used for early warning. Also, we suggested AMI and PVT could predicting genetically the level of four proteins (Dkk-1, PDGF_subunit_B, RAGE and SPON1) and three (IL-1ra, MCP-1 and TIE2) that may be used for auxiliary diagnosis, separately. Nevertheless, there were several limitations. First, the MR analysis was based on European population. Second, the 90 CVPs were taken from the blood, not the myocardium. Third, we did not explore the underlying mechanism in detail, although we found that LEP was a potential mediator in the path from AMI to PVT. Last, the one-sample MR was used for analyzing casual effect between AMI and PVT, but two-sample MR was applied for others.

## Conclusions

In the study, we comprehensively explored the causal effects among AMI, 90 CVPs and PVT. There were three positive (ESM-1, GAL and PRL) and four negative casual effects (FAS, LEP, TNF-R1 and TNF-R2) between genetic liability of AMI on CVPs. There were two positive (IL-27 and MMP-12) and two negative (LEP and TF) casual effects between genetic liability of CVPs on PVT. The AMI played a positive casual effect on PVT mediating by LEP with 5.71%. In addition, we identified two positive (Dkk-1 and PDGF_subunit_B) and two negative (RAGE and SPON1) casual effects between genetic liability of CVPs on AMI, and thee positive (IL-1ra, MCP-1 and TIE2) casual effects between genetic liability of CVPs on PVT.

## Data Availability

Please contact the corresponding author if necessary.

## Abbreviations

AMI: Acute myocardial infarction
PVT: Paroxysmal ventricular tachycardia
CVP: Cardiovascular proteins
MR: Mendelian randomization
UVMR: Univariable Mendelian randomization
IVW: Inverse variance weighted
OR: Odds ratio
CI: Confidence interval
FDR: False discovery rate
AGRP: Agouti-related protein
ESM: 1 Endocan
FAS: Tumor necrosis factor receptor superfamily member 6
GAL: Galanin
LEP: Leptin
PRL: Prolactin
TNF-R1: Tumor necrosis factor receptor 1
TNF-R2: Tumor necrosis factor receptor 2
IL-27: Interleukin-27
MMP-12: Matrix metalloproteinase-12
TF: Tissue factor
Dkk-1: Dickkopf-related protein 1 PDGF_subunit_B Platelet-derived growth factor subunit B
RAGE: Receptor for advanced glycosylation end products
SCF: Stem cell factor
SPON1: Spondin-1
IL-1ra: Interleukin-1 receptor antagonist protein
MCP-1: Monocyte chemotactic protein 1
TIE2: Angiopoietin-1 receptor

## Acknowledgement

The authors thank the investigators of the original studies for sharing the GWAS summary statistics.

## Data and data sharing

Please contact the corresponding author if necessary.

